# Effectiveness and safety of acupuncture for insulin resistance in women with polycystic ovary syndrome: A systematic review and meta-analysis

**DOI:** 10.1101/2022.01.31.22270217

**Authors:** Yu Liu, Hua-ying Fan, Jin-qun Hu, Ming Chen, Jiao Chen

## Abstract

**Objective:** To perform a systematic review and meta-analysis of randomized controlled trials (RCTs) to evaluate acupuncture’s clinical effect of on insulin resistance (IR) in women with polycystic ovary syndrome (PCOS).

**Methods:** PubMed, Cochrane Library, Embase databases, and Chinese databases, including China National Knowledge Infrastructure (CNKI), Technology Journal Database (VIP), and Wanfang Database, were searched without language restrictions from inception to 20 December 2021. Only RCTs in which acupuncture had been examined as the sole or adjunctive PCOS-IR treatment were included. Additionally, only studies in Chinese databases that had been published in core journals of Peking University were included. Our primary endpoint was homeostasis model assessment of insulin resistance (HOMA-IR). The secondary outcomes were fasting blood glucose (FBG), 2-h postprandial blood glucose (2h-PBG), fasting insulin (FINS), body mass index (BMI), and adverse events. A random-effects model enabled reporting of differences between groups as mean differences, thus minimizing the effects of uncertainty associated with inter-study variability on the effects of different interventions.

**Results:** Our analysis included seven eligible RCTs (N=728 participants). Compared with other treatments, acupuncture therapy yielded a greater mean reduction in BMI (−1.21; 95% CI, −2.41 to −0.02; P=0.05). No significant differences existed between acupuncture and other studied treatments for changes in HOMA-IR (−0.33; 95% CI, −0.87 to 0.22; P>0.05), FBG (−0.43; 95% CI, −0.88 to 0.03; P=0.07), 2h-PBG (−0.40;95% CI, −0.90 to 0.10; P>0.05), and FINS (−0.65; 95% CI, −2.18 to 0.89; P>0.05). Furthermore, compared with medication alone, a combination of acupuncture and medication yielded a mean reduction in HOMA-IR of −0.63 (95% CI, −1.12 to −0.14; P=0.01) and BMI of −1.36 (95% CI, −2.07 to −0.66; P<0.01).

**Conclusion:** Although acupuncture is not more effective than metformin, the former could be an adjuvant strategy for improving PCOS-IR. Further large-scale, long-term RCTs with strict methodological standards are justified.

## 1 Introduction

Polycystic ovary syndrome (PCOS), a complex endocrine and metabolic disorder, is characterized by androgen excess (hirsutism and/or hyperandrogenemia) and ovarian dysfunction (oligo-ovulation and/or polycystic ovarian morphology). To date, its pathogenesis remains unclear. However, insulin resistance (IR) is considered the primary pathological basis for the associated reproductive dysfunction. The prevalence of IR in clamp studies in women with PCOS diagnosed on the basis of the Rotterdam criteria and using age-appropriate lean healthy control women is reportedly 75% of lean and 95% of overweight women (weight status according to WHO criteria) [1]. IR both promotes and interacts with hyperandrogenemia, which affects the function of the hypothalamic–pituitary–ovarian axis and causes abnormal follicular development [2].

Additionally, regardless of age, gestational diabetes mellitus, impaired glucose tolerance, and type 2 diabetes are all significantly more prevalent in patients with PCOS. Some studies have found that improving IR and reducing hyperinsulinemia can reduce systemic androgen concentrations and improve some characteristics of PCOS [3]. Metformin is often recommended to adult women or adolescents with PCOS or women with BMI>25 kg/m^2^ for management of weight and metabolic disorders [4]. However, the mechanism underlying metformin’s effects on PCOS-IR is not yet fully understood and it causes adverse effects, particularly diarrhea and nausea, in up to 25% of patients [5]. Thus, metformin has limitations related to adverse effects and patient compliance. There is therefore a need for inexpensive and easily administered treatments with few adverse effects. Correlation meta-analysis has shown that acupuncture has a beneficial effect on IR and fewer adverse reactions than other treatments [6,7]. A recent meta-analysis also found that acupuncture can improve IR in patients with PCOS [8]. However, this evidence is inadequate because of the poor quality and methodology of the included studies. In addition, these authors did not determine whether acupuncture has advantages over metformin in the treatment of PCOS-IR. In the present study, we systematically reviewed data from recent studies, aiming to provide clarity concerning the role of acupuncture in treatment of PCOS-IR, especially compared with metformin.

In this systematic review of relevant randomized controlled trials, we aimed to evaluate the efficacy and safety of acupuncture in improving PCOS-IR by assessing the following key outcomes: homeostasis model assessment of insulin resistance (HOMA-IR), fasting blood glucose (FBG), 2-h postprandial blood glucose (2h-PBG), fasting insulin (FINS), body mass index (BMI), and adverse events.

## 2 Methods

This analysis was performed strictly in accordance with the PRISMA statement [9] (see S1 File). Additionally, this review was registered with PROSPERO at http://www.crd.york.ac.uk/PROSPERO(CRD42021285851).

### 2.1 Search strategy

PubMed, Cochrane Library, and Embase databases were searched without language restrictions from inception to 20 December 2021. We also searched Chinese databases, including the China National Knowledge Infrastructure (CNKI), Technology Journal (VIP), and Wanfang databases. Additionally, only studies in Chinese databases that had been published in core journals of Peking University (such as *Chinese Acupuncture and Moxibustion*) were included. The search terms consisted of four parts: acupuncture (acupuncture, electroacupuncture, and manual acupuncture), insulin resistance (insulin resistance and insulin sensitivity), polycystic ovary syndrome (polycystic ovary syndrome, polycystic ovarian syndrome and Stein–Leventhal syndrome), and randomized controlled trial. Details of the search strategies are presented in S2 File.

Additionally, ClinicalTrials.gov and the Chinese Clinical Trial Register were searched to identify ongoing or recently completed studies.

### 2.2 Inclusion criteria

#### 2.2.1 Types of study

All randomized controlled trials (RCTs) involving use of acupuncture for treating PCOS-IR were included.

#### 2.2.2 Types of participant

Patients diagnosed with PCOS-IR or PCOS patients with HOMA-IR≥ 2.14 [10], regardless of race, or educational and economic status, were included.

#### 2.2.3 Types of intervention

Interventions in the experimental group comprised acupuncture or electroacupuncture either combined with medication or not. Controlled interventions with sham acupuncture or metformin were included.

#### 2.2.4 Types of outcome measure

Studies that reported at least one clinical outcome related to PCOS-IR were included. The primary outcome was HOMA-IR. The secondary outcomes were FBG, 2h-PBG, FINS, BMI, and adverse events.

### 2.3 Exclusion criteria

#### 2.3.1 Types of study

Nonrandomized controlled trials, randomized crossover trials, reviews, case reports, protocols, experimental animal articles were excluded.

#### 2.3.2 Participants

Participants with serious physical or mental disease were excluded.

#### 2.3.3 Types of intervention

We did not include trials in which non-penetrating acupuncture (such as via laser stimulation, acupressure, or transcutaneous electrical nerve stimulation) was used or in which other traditional Chinese medicine was administered to the experimental group. RCTs that compared different forms of acupuncture or herbal medicine were also excluded.

### 2.4 Study identification and data extraction

Two independent appraisers (Yu Liu and Jin-qun Hu) assessed the eligibility of the searched articles in accordance with the above criteria. The following data were extracted independently from all included studies by two reviewers (Yu Liu and Ming Chen): number of participants, age, interventions, intervention duration, change in HOMA-IR (mean [SD]); change in FBG (mean [SD]); change in 2h-PBG (mean [SD]); change in FINS (mean [SD]); change in BMI (mean [SD]), and number of participants with any adverse events. Any disagreements regarding study identification and data extraction were resolved by discussion and adjudication with a third investigator (Hua-ying Fan).

### 2.5 Quality assessment

Two reviewers (Yu Liu and Jin-qun Hu) independently assessed the risk of bias of RCTs using the Cochrane Collaboration’s “risk of bias” tool, which is based on the following six separate domains: random sequence generation (selection bias), allocation concealment (selection bias), blinding of participants and personnel (performance bias), blinding of outcome assessment (detection bias), incomplete outcome data (attrition bias), selective reporting (reporting bias), and other bias. The assessments were categorized into three levels of bias: low risk, high risk, or unclear risk. For example, if the investigators described a random component in the sequence generation process, such as using a computer random number generator or coin tossing, “low risk” would be allocated for the domain of random sequence generation. Disagreements were resolved by a third investigator (Hua-ying Fan).

We also used Grading of Recommendations Assessment, Development, and Evaluation (GRADE) to assess the quality of evidence, using GRADEpro GOT online software.

### 2.6 Data synthesis

We assessed the effect and safety of acupuncture for treating PCOS-IR on the basis of six outcomes: HOMA-IR, FBG, 2h-PBG, FINS, incidence of adverse events, and BMI. We analyzed HOMA-IR, FBG, 2h-PBG, FINS, and BMI as continuous variables and have reported absolute differences between arithmetic means before and after interventions. Adverse events were treated as a categorical variable and the risk ratio calculated.

We calculated pooled estimates of the mean differences (MD) in HOMA-IR, FBG, 2h-PBG, FINS, and BMI between intervention groups using a random-effects model (DerSimonian–Laird method) to minimize additional uncertainty associated with inter-study variability regarding the effects of different interventions. We also calculated pooled risk ratio estimates for categorical outcomes with a random-effects model (DerSimonian–Laird method).

We used the Cochran Q test to assess heterogeneity between studies. P values less than 0.1 were considered to denote statistical significance. We also did *I*^2^ testing to assess the magnitude of heterogeneity between studies, values greater than 50% being regarded as indicative of moderate-to-high heterogeneity. If necessary, subgroup analyses according to type of intervention were conducted. We used RevMan (version 5.4) for all statistical analyses.

## 3. Results

### 3.1 Search results

A flow chart of study selection is shown in Fig 1. Our preliminary search found a total of 723 articles. Removing duplicate articles and reading the title and summary filters left 62 full-text articles, 53 of which were excluded because they did not meet the inclusion criteria. Two more articles were excluded because their data were unavailable and we received no responses from the corresponding authors. Finally, seven studies including 728 patients were included in this review.

**Fig 1.**
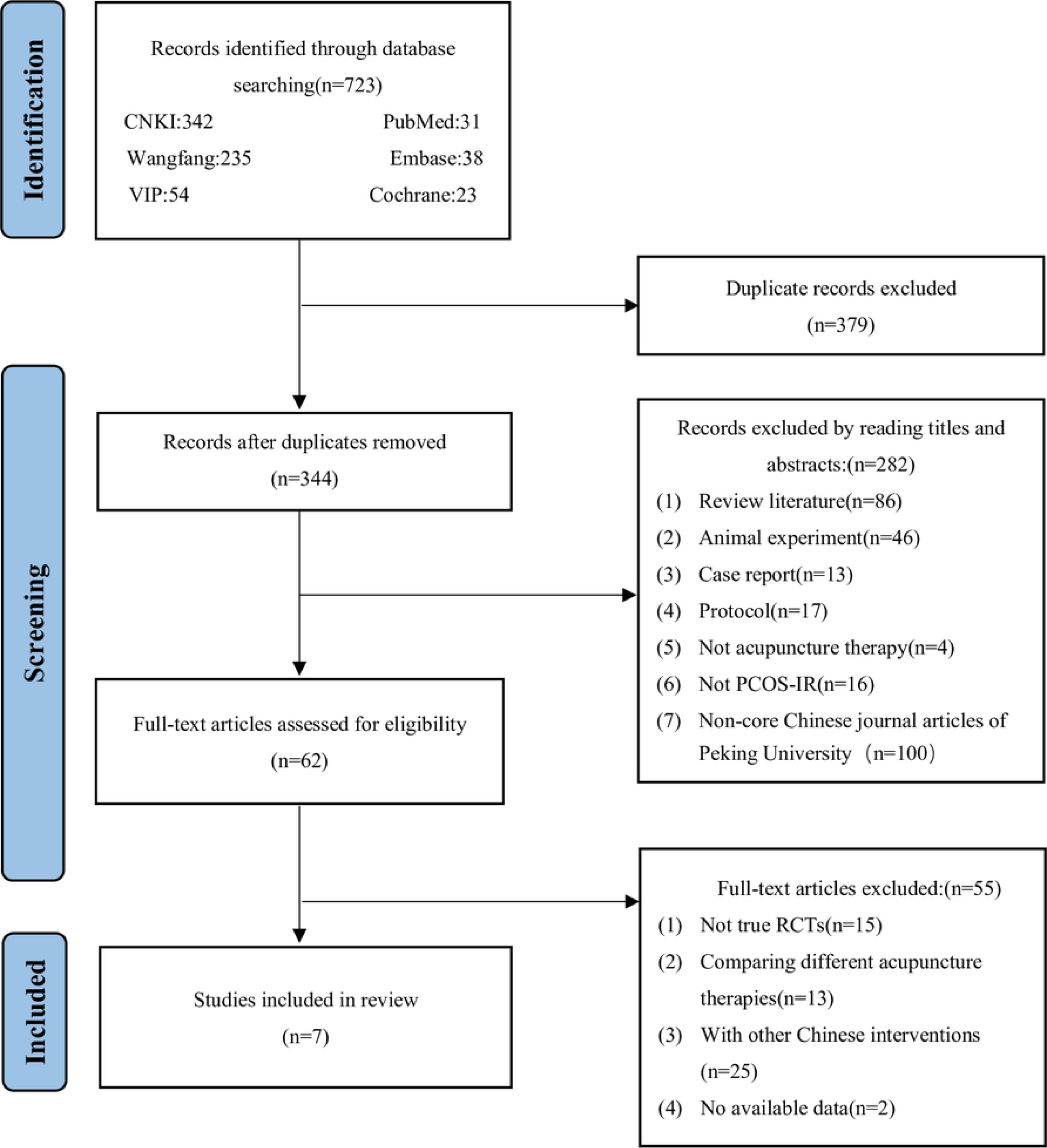
Literature screening process and results.

### 3.2 Description of included studies

The seven eligible RCTs including 728 patients were conducted in China. Three of them were listed in English databases and four in Chinese databases. All participants had been diagnosed with PCOS in accordance with the Rotterdam criteria and had HOMA-IR greater than 2.14, enabling diagnosis of IR [10]. The patients had a mean baseline HOMA-IR of 4.02 (range 2.37–5.06) and mean baseline BMI of 27.18 kg/m^2^ (22.4–29.35). Electroacupuncture was compared with sham acupuncture in two studies [11,12], acupuncture combined with medication was compared with medication alone in two studies [13,14], and acupuncture was compared with metformin in three studies [15–17]. Table 1 summarizes the basic characteristics of these studies.

**Table 1.** Characteristics of included studies.

| Study ID | Sample<br>size<br>(TG/CG) | Intervention |  | Duration of<br>intervention | Mean<br>age<br>(years) | Mean<br>BMI<br>(kg/m<sup>2</sup>) | Mean<br>HOMA-IR | Outcomes |
| --- | --- | --- | --- | --- | --- | --- | --- | --- |
|  |  | TG | CG |  |  |  |  |  |
| Dong,2021[11] | 27/27 | EA | SA | 16 weeks | 22.8 | 22.4 | 2.37 | ①②④⑤ |
| Gu,2019[12] | 39/39 | EA | SA | 32 times | 27.8 | 27.6 | 5.06 | ①②③④⑤⑥ |
| Li,2014[13] | 53/51 | A+M | SA+M | 6 months | 26.1 | 29.35 | 3.75 | ①②④⑤⑥ |
| Mao,2021[14] | 54/54 | EA+M+E | M+E | 3 MC | Not<br>available | 28.32 | 3.83 | ①⑤ |
| Wen,2021[15] | 114/114 | EA+P | SA+M | 4 months | 27.7 | 26.15 | 4.47 | ①②④⑤⑥ |
| Yu,2020[16] | 36/34 | EA | M | 3 months | 30.5 | Not<br>available | 4.81 | ①⑥ |
| Zheng,2013[17] | 43/43 | A | M | 6 months | 25.7 | 29.25 | 3.85 | ①②③④⑤⑥ |

A, acupuncture; CG, control group; E, ethinylestradiol and cyproterone acetate tablets; EA, electroacupuncture; M, metformin; MC, menstrual cycle; P, placebo; SA, sham acupuncture; TG, treatment group; ①, HOMA-IR; ②, FBG; ③, 2h-PBG; ④, FINS; ⑤, BMI; ⑥, adverse events.

### 3.3 Assessment of risk of bias

The participants were randomized using a random number table or computer program in all seven studies. Assessment of allocation was described in three articles [11,15,17]. For blinding, three trials [11,13,15] were classified as “low risk” and the others as “unclear risk”. Three articles [11,15,16] stated explicitly that third-party researchers processed the data. Four studies[11,12,15,16] reported loss to follow-up. There was attrition bias in two of these studies [11,15] as a result of differences in the proportion of missing outcome data between the experimental and comparison groups. The other three trials had a low risk of attrition bias, having reported that no participants dropped out or were excluded from the primary analysis. Details of the assessments are shown in Figs 2 and 3.

**Fig 2.**
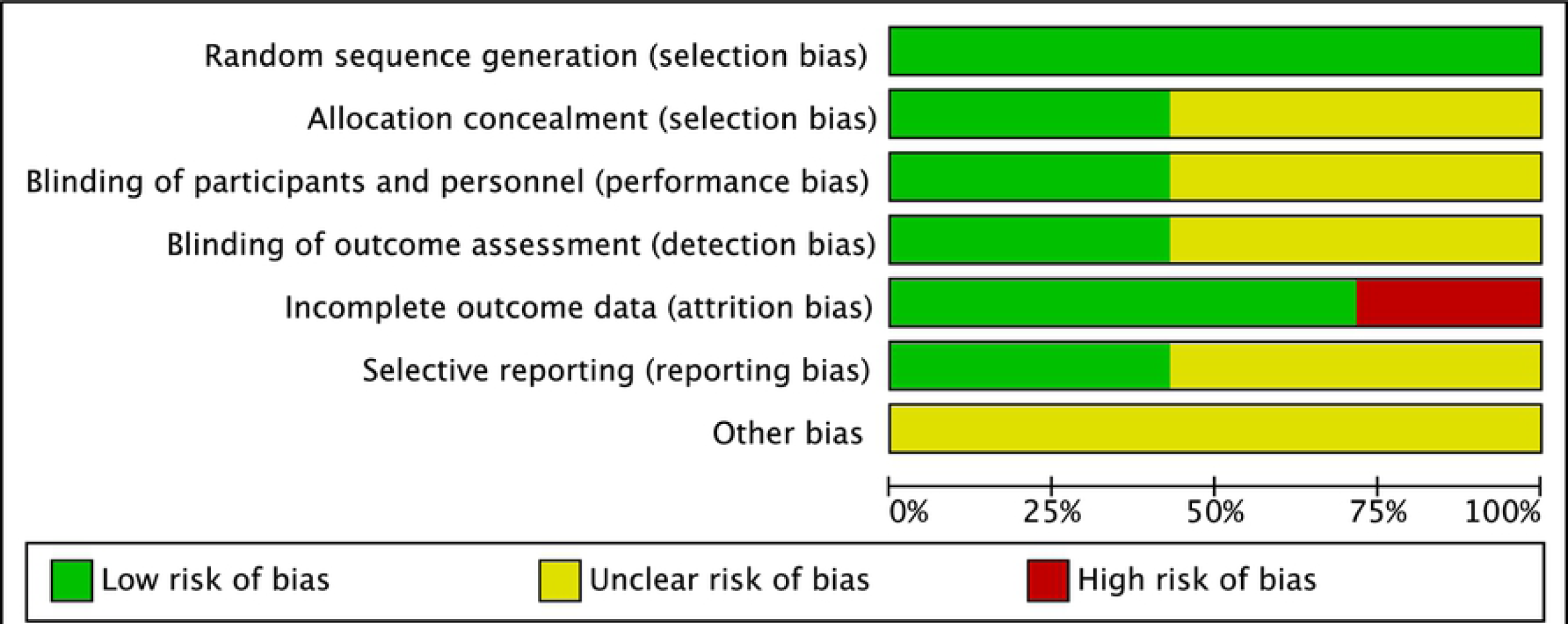
Assessment of risk biases of the included studies.

**Fig 3.**
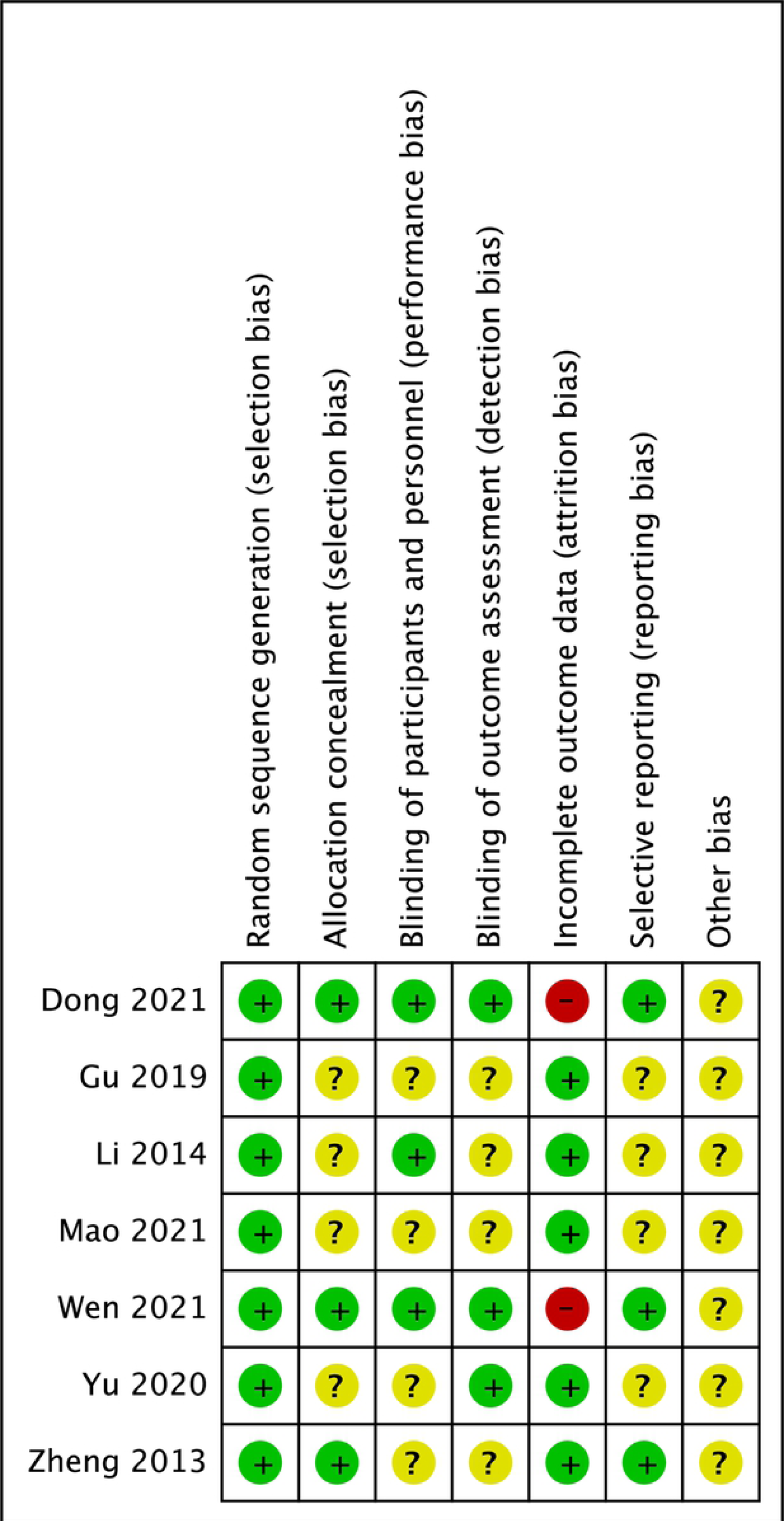
Assessment of risk biases of the included studies.

### 3.4 Meta-analysis of primary outcomes

A pooled meta-analysis analysis of the seven trials found no statistically significant differences in changes in HOMA-IR (MD, −0.33; 95% CI, −0.87 to 0.22; P=0.24; Fig 4) between the two groups, with statistically significant heterogeneity (P<0.1; *I*^2^=82%).

**Fig 4.**
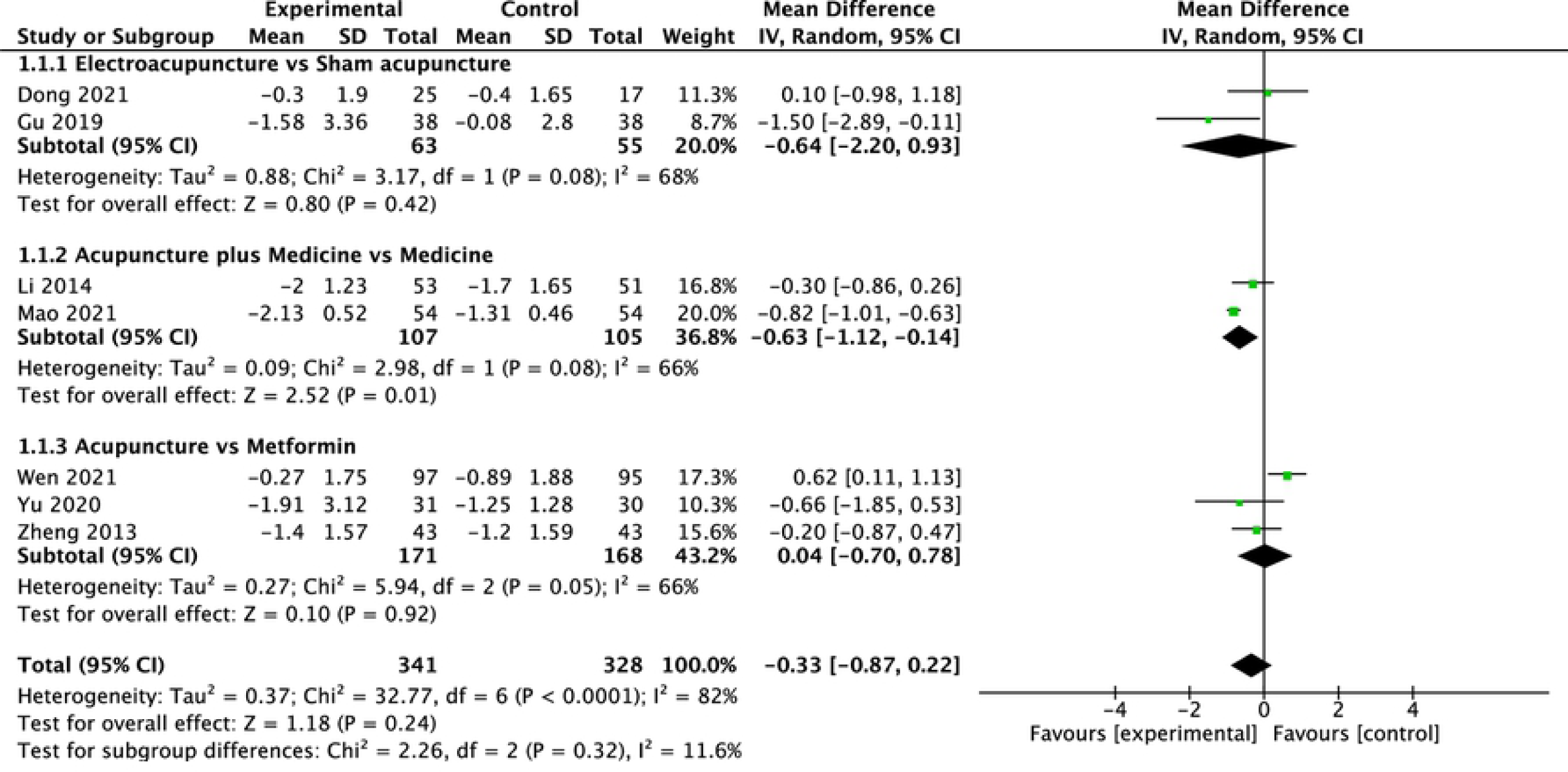
Comparison of changes in HOMA-IR according to intervention.

To further analyze heterogeneity, we performed subgroup analysis of different interventions to more accurately examine the effects of acupuncture. Sensitivity analysis comparing acupuncture treatment with metformin or sham acupuncture identified no significant differences (MD, 0.04; 95% CI, −0.7 to 0.78; P=0.92; Fig 4) (MD, −0.64; 95% CI, −2.2 to 0.93; P=0.42; Fig 4). However, comparison of a combination of acupuncture and medication with medication alone showed a greater mean reduction in HOMA-IR with the former (MD, −0.63; 95% CI, −1.12 to −0.14; P=0.01; Fig 4).

### 3.5 Meta-analysis of secondary outcomes

#### 3.5.1 FBG

Pooled analysis of the five studies that assessed changes in participants’ FBG after treatment showed a trend to improvement in FBG that was not statistically significant (MD, −0.43; 95% CI, −0.88 to 0.03; P=0.07; Fig 5), with statistically significant heterogeneity (P<0.1; *I*^2^=86%).

**Fig 5.**
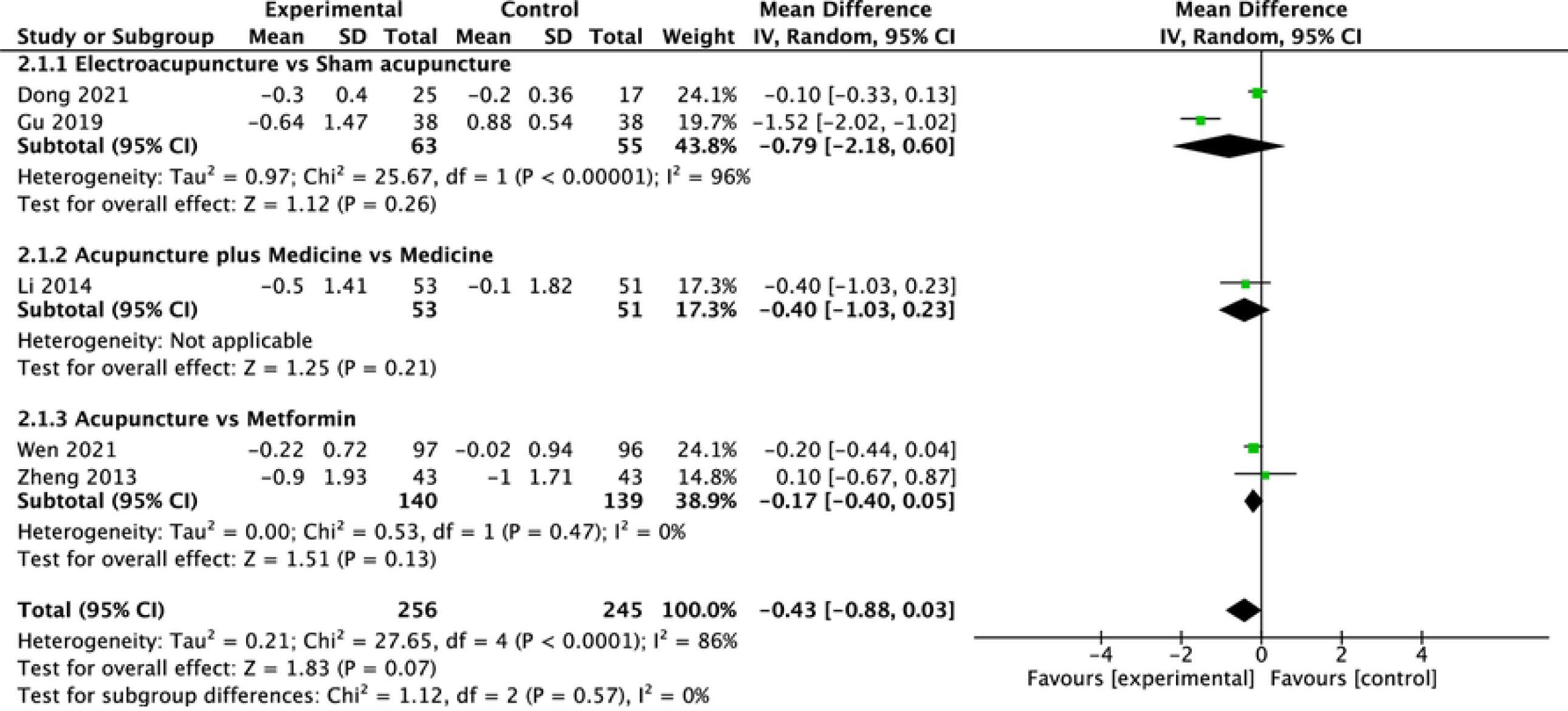
Comparison of changes in FBG according to intervention.

Forest plots generated in further sensitivity analysis showed no significant difference between acupuncture and metformin (MD, −0.17; 95% CI, −0.4 to 0.05; P=0.13; Fig 5). Additionally, combination treatment showed no benefit over medication alone (MD, −0.4; 95% CI, −1.03 to 0.23; P=0.21; Fig 5).

#### 3.5.2 2h-PBG

Two studies (N=162 participants) assessed changes in 2h-PBG at the end of the intervention. Pooling the data of these studies showed no significant difference in mean reduction of 2h-PBG between acupuncture and other treatments (MD, −0.40; 95% CI, −0.90 to 0.10; P=0.12; Fig 6), without between-study heterogeneity.

**Fig 6.**
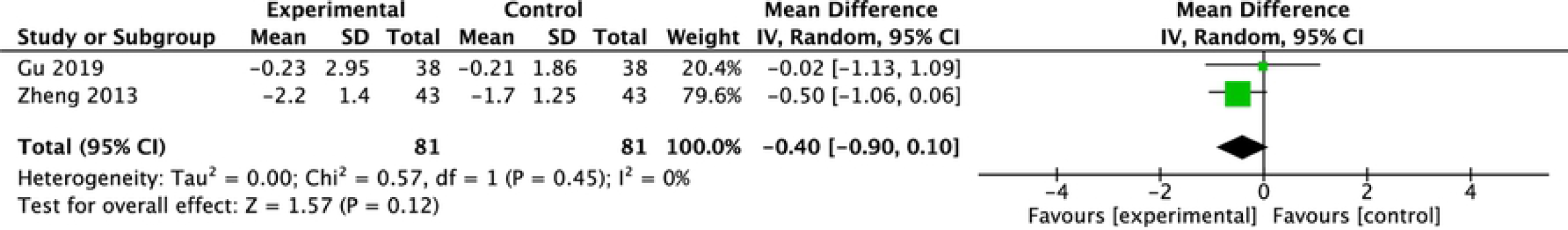
Comparison of changes in 2h-PBG according to intervention.

#### 3.5.3 FINS

Five studies (N=502 participants) assessed changes in participants’ FINS after treatment. Pooled analysis of these studies’ findings showed that acupuncture was not more effective than other treatments regarding mean reduction in FINS (MD, −0.65; 95% CI, −2.18 to 0.89; P=0.41; Fig 7), with no significant between-study heterogeneity (P>0.1; *I*^2^=37%).

**Fig 7.**
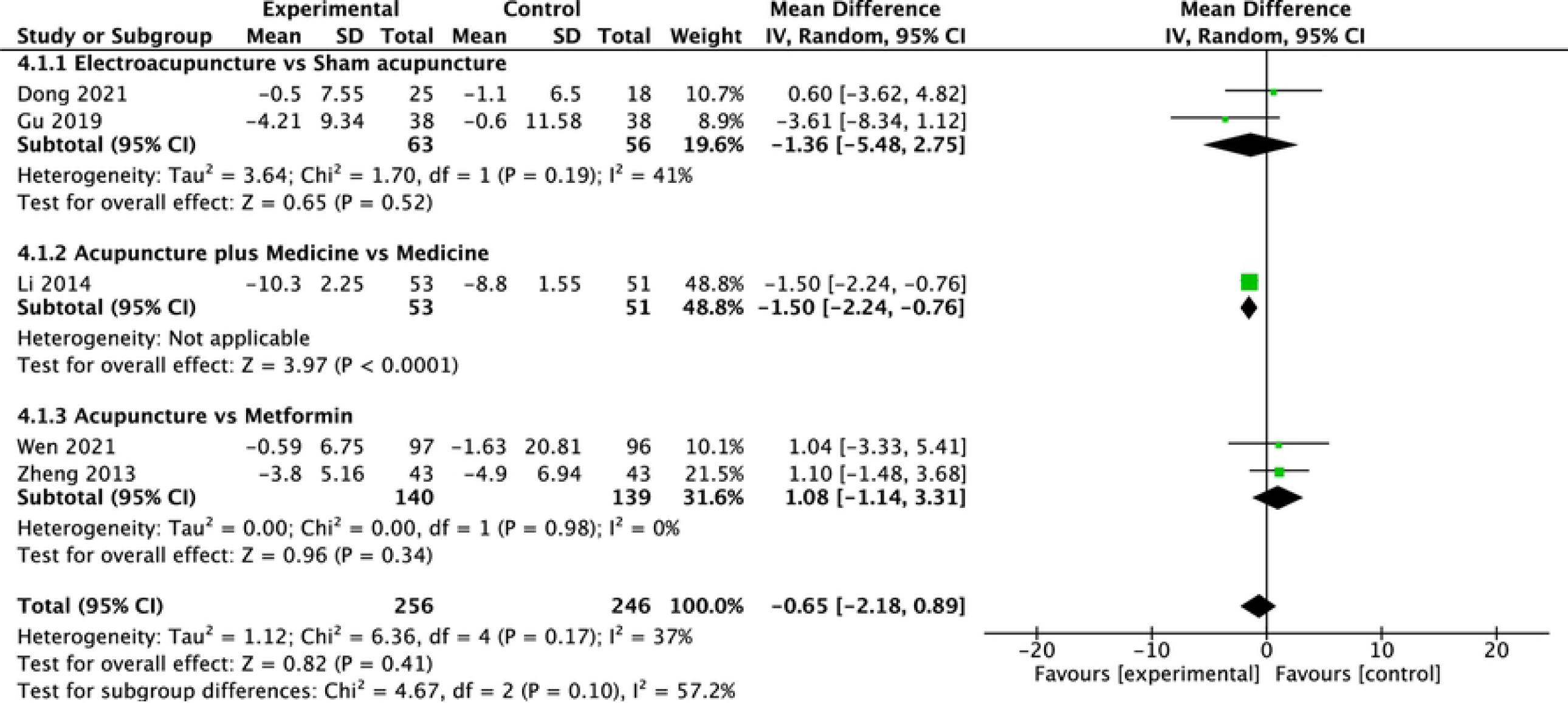
Comparison of changes in FINS according to intervention.

#### 3.5.4 BMI

Six studies assessed changes in participants’ BMI after treatment. Analysis of pooled data of these studies showed that acupuncture was associated with a greater reduction in weight than other treatment (MD, −1.21; 95% CI, −2.41 to −0.02; P=0.05; Fig 8), with statistically significant heterogeneity (P<0.1; *I*^2^=88%).

**Fig 8.**
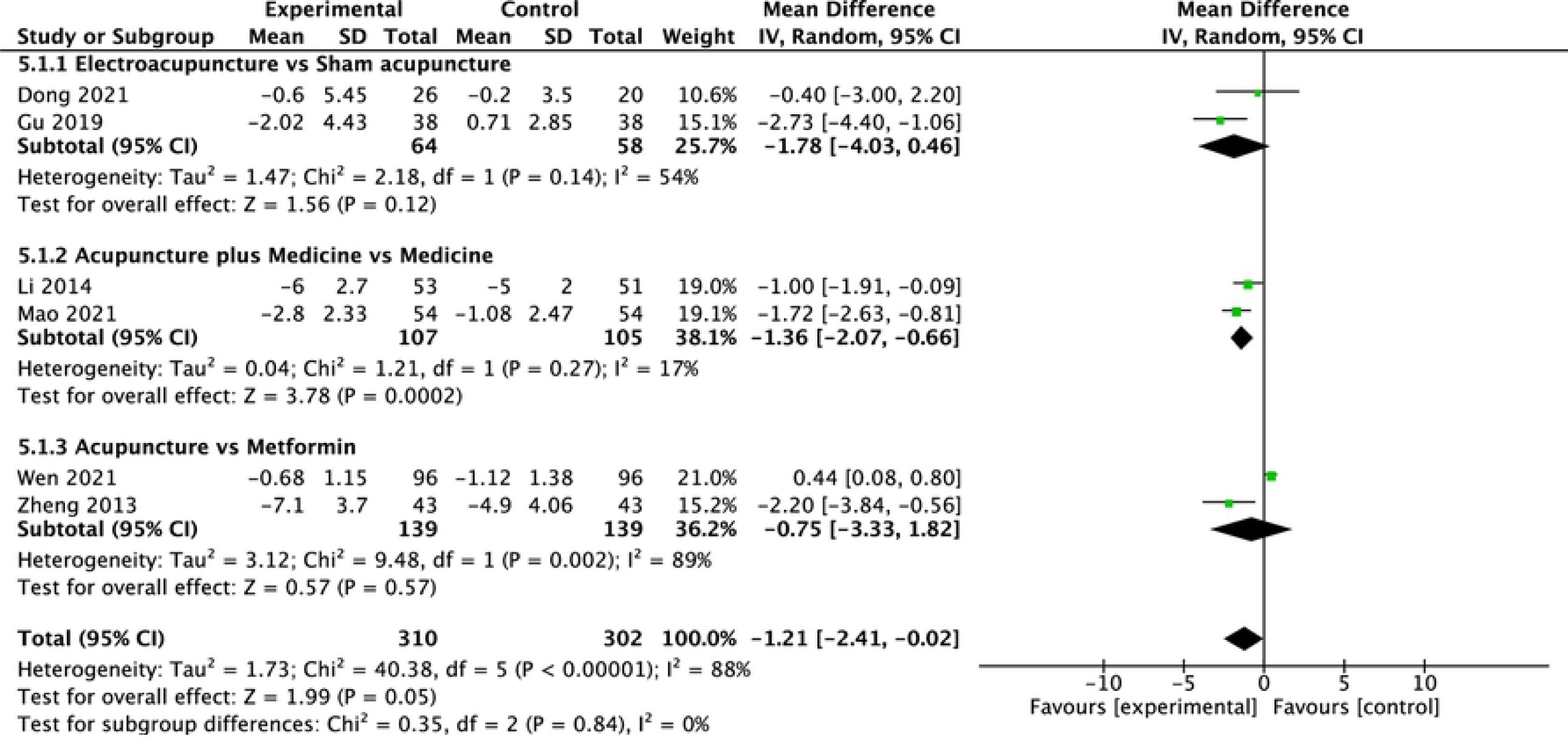
Comparison of changes in BMI according to intervention.

Sensitivity analysis revealed no statistically significant difference in changes in BMI between acupuncture and metformin (MD, −0.75; 95% CI, −3.33 to 1.82; P=0.57; Fig 8). However, a combination of acupuncture and medication achieved a greater mean reduction in BMI than did medication alone (MD, −1.36; 95% CI, −2.07 to −0.66; P<0.01; Fig 8).

#### 3.5.5 Adverse events

Five of the studies reported adverse events. Three of these studies compared acupuncture with metformin, one compared combined treatment with acupuncture and medication with medication alone and one compared electroacupuncture with sham acupuncture. One [16] reported 29.4% cases of nausea, emesis, diarrhea, and loss of appetite in the control group and 11.1% of subcutaneous bruising and hematoma in the experimental group. One [15] reported 87.7% adverse gastrointestinal events in the control group and 14.9% cases of subcutaneous bruising and hematoma in the experimental group. One [17] reported 48.8% adverse effects in the control group and none in the experimental group. One [13] reported 43.1% cases of nausea, emesis, diarrhea, and loss of appetite in the control group and 34% adverse events in the experimental group. Most adverse events were digestive tract reactions or subcutaneous bruising and hematoma and resolved quickly. There were no distressing symptoms or liver or kidney injury. Table 2 displays the reported details.

**Table 2.** Reported adverse events.

| Study ID | Sample size |  | Intervention |  | Nausea, emesis, diarrhea, and loss of appetite |  | Subcutaneous bruising and hematoma |  |
| --- | --- | --- | --- | --- | --- | --- | --- | --- |
|  | TG | CG | TG | CG | TG | CG | TG | CG |
| Gu,2019[12] | 39 | 39 | EA | SA | - | - | 0 | 0 |
| Li,2014[13] | 53 | 51 | A+M | SA+M | 18 | 22 | - | - |
| Wen,2021[15] | 114 | 114 | EA+P | SA+M | - | 100 | 17 | - |
| Yu,2020[16] | 36 | 34 | EA | M | - | 10 | 4 | - |
| Zheng,2013[17] | 43 | 43 | A | M | - | 21 | 0 | - |

A, acupuncture; CG, control group; EA, electroacupuncture; M, metformin; P, placebo; SA, sham acupuncture; TG, treatment group.

### 3.6 Quality of evidence

The quality of evidence assessed using the GRADE system varied from very low to low. According to this system, FINS had a low evidence level because the means of assessing allocation was not described, and the upper or lower confidence limits crossed an effect size of 0.5 in both directions. Further, HOMA-IR, FBG, 2h-PBG, and BMI had very low evidence levels. Details of assessment are shown in Table 3.

**Table 3.**
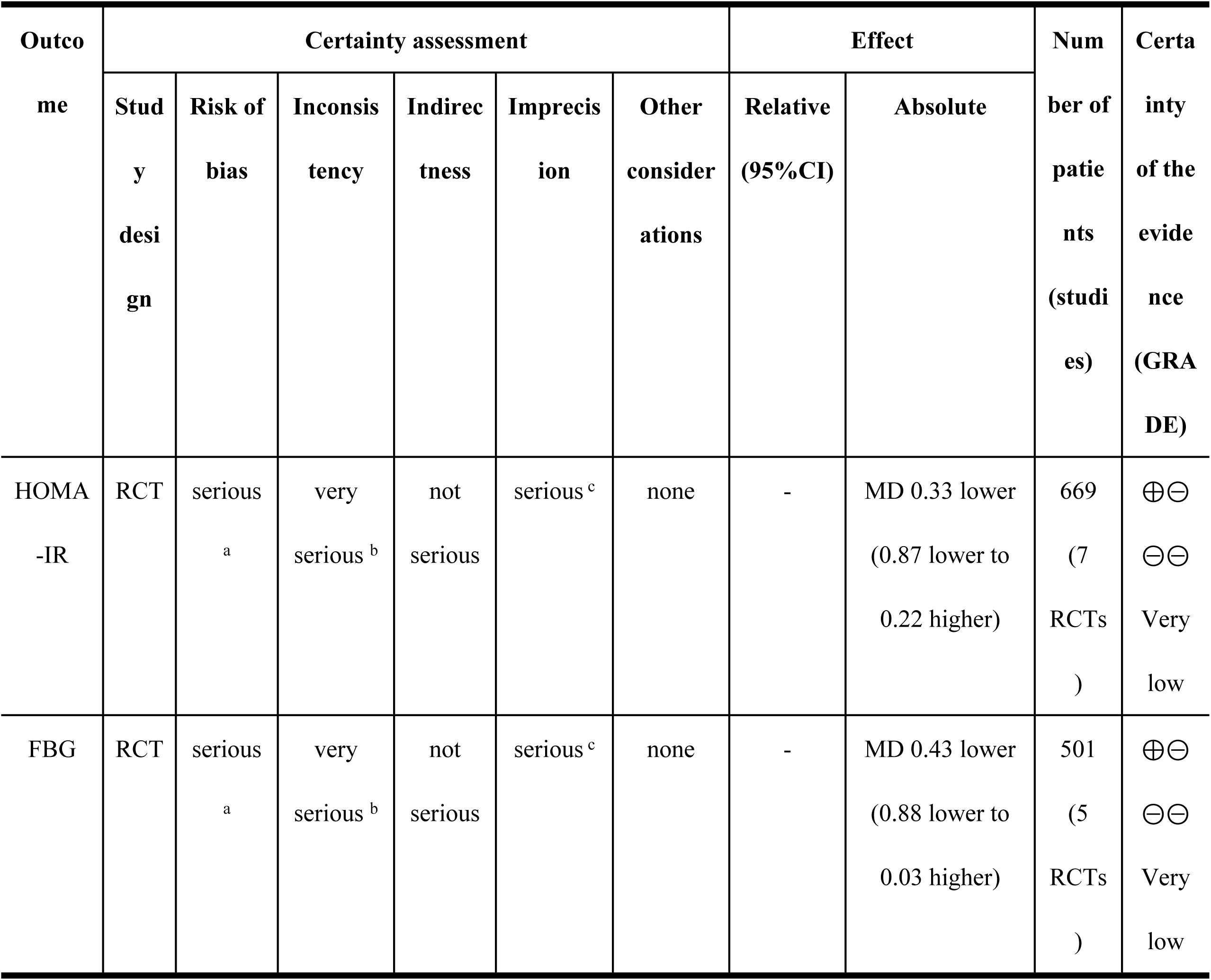
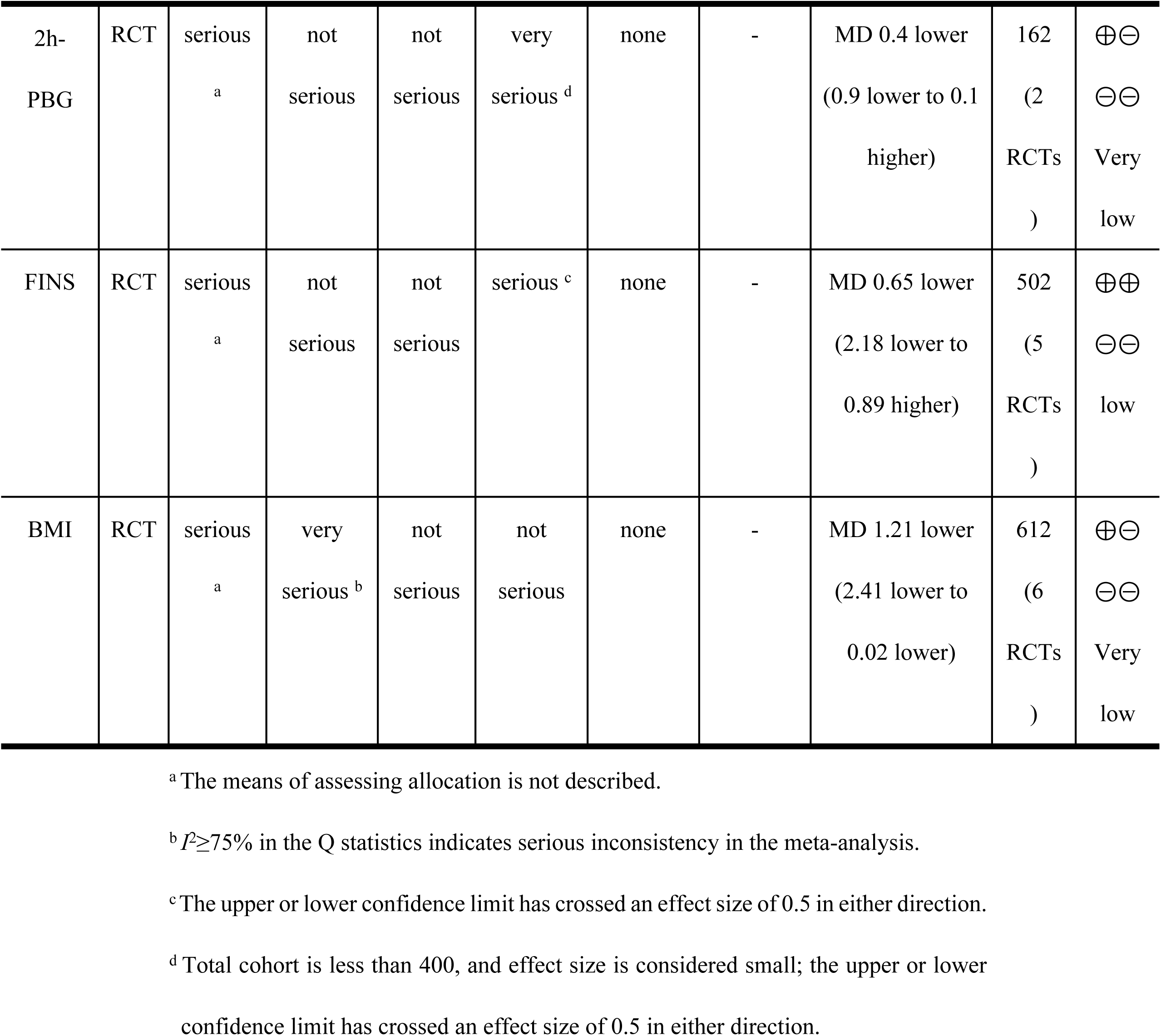
Assessment according to GRADE system.

## Discussion

### Main findings

The purpose of this review was to evaluate the effectiveness of acupuncture for IR in patients with PCOS. We found acupuncture to be significantly associated with greater loss of body weight (based on very low-certainty evidence) than other treatments.

However, we identified no significant differences for the other studied outcomes, namely FINS (based on low-certainty evidence), and HOMA-IR, FBG, and 2h-PBG (all based on very low-certainty evidence). Although acupuncture is not more effective than metformin in improving IR, it does not cause nausea, emesis, or diarrhea, and any subcutaneous hematoma will resolve quickly. Furthermore, we found that, compared with medication alone, a combination of acupuncture and medication yields improvements in HOMA-IR and BMI with fewer gastrointestinal adverse events. Therefore, our findings indicate that acupuncture is a promising adjuvant strategy for treating PCOS-IR.

The cause of PCOS is unknown. However, it is strongly associated with IR and the risk of type 2 diabetes in PCOS patients is 5–10 times higher than that in the general population [18]. Clinical and experimental evidence has shown that acupuncture is an acceptable, adverse effect-free alternative or complement to pharmacological induction of ovulation in women with PCOS and may also relieve other symptoms [19]. There is also evidence that acupuncture can be an insulin sensitizer and may therefore contribute to controlling obesity and type 2 diabetes [20,21]. Acupuncture ameliorates IR through enhancing autophagy [22], affecting insulin receptor signal transduction, and increased expression of insulin receptor substrate in the endometrium in a PCOS-like rat model [23,24].

### Limitations

First, most studies examined were small and some of them described the specifics of assessing allocation and blinding poorly. Their quality was therefore mediocre. Besides, according to the GRADE system, only one outcome was based on low-certainty evidence, the other four outcomes being based on very low-certainty evidence. Second, these studies lack follow-up data, hindering assessment of the long-term efficacy of acupuncture for PCOS-IR.

Finally, there was heterogeneity in some outcomes. We accordingly performed subgroup analyses; however, the likelihood of false negative and false positive findings regarding significance increase rapidly with increasing numbers of subgroup analyses. Of note, metabolic syndrome has varying clinical manifestations and biochemical characteristics, age distribution, and abnormal glucose metabolism. Further, there were differences in the details of the interventions, severity of disease, dosage of metformin, and acupuncture protocols between studies.

### Implications for further clinical research

Being a safe and simple treatment modality, acupuncture could be a good alternative or adjuvant therapy for PCOS-IR, especially for overweight patients. Because it has fewer adverse effects than metformin, patient acceptance and compliance may be higher. Regardless of age, the prevalences of gestational diabetes mellitus, impaired glucose tolerance, and type 2 diabetes are significantly greater in individuals with PCOS than in those without it. The risk is not related to obesity but is increased by obesity [4]. Successful treatment of obesity therefore reduces IR [25]. It is gratifying that acupuncture is significantly more effective at achieving weight loss than other treatments. Unfortunately, the long-term effects of acupuncture are still unknown. Additional long-term follow-up studies are needed to clarify the role of acupuncture in the treatment of PCOS-IR.

In addition, some comparative studies of acupuncture and sham acupuncture have found that these two interventions achieve equivalent results. A review of the designs of these studies revealed that sham acupuncture involved needling non-acupuncture or irrelevant acupuncture points, or superficial needling. However, insertion of a needle into a recognized acupuncture or non-acupuncture point can produce a physiological effect, partly by activating the pain inhibiting system in the spinal cord and diffuse noxious inhibitory controls [26–29]. In addition, sham acupuncture applied at non-acupuncture points may serve as an active control because acupoint areas can be enlarged by increased expression of nociceptive substances in individuals with painful conditions [30]. It has therefore been suggested that more research into applying non-penetrating sham acupuncture at non-acupuncture points is necessary [31]. This would avoid segmental analgesia and minimize any physiological effect in the sham acupuncture group.

## Conclusions

This review of seven RCTs shows that acupuncture can reduce body weight significantly in individuals with PCOS-IR and that, while not being more effective than metformin, it has fewer adverse effects. Additionally, combination treatment with acupuncture and medication more effectively improves HOMA-IR and BMI, with less gastrointestinal adverse events, than medication alone. Acupuncture may be an effective adjuvant strategy for improving PCOS-IR. However, the evidence for this conclusion is based on only a few studies with obvious limitations related to both sample size and methodology. Further large-scale, long-term RCTs with strict methodological standards are justified.

## Data Availability

All relevant data are within the manuscript and its Supporting Information files.

## Acknowledgment

We thank Dr Trish Reynolds, MBBS, FRACP, from Liwen Bianji (Edanz) (www.liwenbianji.cn/), for editing the English text of a draft of this manuscript.

## Supporting information

**S1 File. PRISMA Checklist**

(DOC)

**S2 File. Search strategies used in English databases**

(DOC)

